# Immunometabolic Alterations in Post-Traumatic Stress Disorder

**DOI:** 10.64898/2026.03.20.26348619

**Authors:** Jelena Brasanac, Linda El-Ahmad, Emma Mollerup, Stefanie Gamradt, Luisa Gruenberg, Daria Shyshko, Victoria Stiglbauer, Kim Zimbalski, Nikola Schoofs, Kathlen Priebe, Felix Wülfing, Simon Guendelman, Tolou Maslahati, Stefanie Koglin, Christian Otte, Isabel Dziobek, Stefan Roepke, Stefan M. Gold

## Abstract

Post-traumatic stress disorder (PTSD) has been linked to various alterations within the immune system, yet the metabolic programming of immune cells remains unexplored. In the current cross-sectional study, we interrogated immunometabolic function by applying cell-specific metabolic flow cytometry, serum biomarker profiling, and targeted gene expression analysis in peripheral blood mononuclear cells from patients with PTSD (N=34) compared with healthy controls (N=32).

PTSD was associated with higher glycolysis- and oxidative pentose phosphate pathway-related markers across adaptive and innate immune cell subsets, as well as elevated circulating interleukin-6. Expression of inflammatory- and stress-related genes was largely comparable between groups.

Together, these data provide preliminary evidence for immunometabolic alterations in PTSD at both cellular and systemic levels. These results could contribute to understanding potential pathophysiological mechanisms and support further investigation of immunometabolism in PTSD.

## 1. Introduction

Post-traumatic stress disorder (PTSD) is a severe, chronic, and heterogeneous psychiatric condition that may develop following exposure to a traumatic event (*Diagnostic and Statistical Manual of Mental Disorders*, 2013). The lifetime prevalence of PTSD in the general population is 3.9% (Koenen et al., 2017). Clinically, PTSD is characterized by intrusive re-experiencing of traumatic memories, avoidance of trauma-related cues, negative alterations in mood and cognition and hyperarousal, including exaggerated threat responses and impaired fear extinction (*Diagnostic and Statistical Manual of Mental Disorders*, 2013).

PTSD is frequently comorbid with other psychiatric disorders (Bryant, 2022; Qassem et al., 2021) as well as physical health conditions involving immune and metabolic dysregulation (Mellon et al., 2018; Michopoulos et al., 2016; Song et al., 2018). For example, links exist between PTSD and inflammatory bowel disease, rheumatoid arthritis, multiple sclerosis, psoriasis, cardiovascular disease, type 2 diabetes, and metabolic syndrome and its components (obesity, insulin resistance, dyslipidemia) (Allgire et al., 2021; Bookwalter et al., 2020; Boscarino, 2004; O’Donnell et al., 2021; O’Donovan et al., 2015; Wilson et al., 2019; Bartoli et al., 2015; Farr et al., 2015; Rosenbaum et al., 2015; Suliman et al., 2016).

It remains unclear whether PTSD drives systemic inflammation (Solomon et al., 2017; Toft et al., 2018), whether immune dysregulation contributes to its etiology (Cohen et al., 2011; Eraly et al., 2014; Pervanidou et al., 2007), or whether this relationship is bidirectional (Sumner et al., 2020). In addition, comorbid metabolic conditions can worsen inflammatory processes, and the metabolic and neuroendocrine alterations observed in PTSD may either predispose individuals to systemic metabolic dysfunction or represent a primary consequence of trauma exposure (Michopoulos et al., 2016). Together, these findings point to a potential contribution of metabolic dysregulation to PTSD, with immune dysfunction as a shared underlying mechanism (Michopoulos et al., 2016).

Over the past decades, growing evidence has highlighted the role of the immune system in PTSD pathophysiology (Hori & Kim, 2019; Michopoulos et al., 2017; Sumner et al., 2020) (for a comprehensive review, see (Katrinli et al., 2022)). Case-control studies, along with follow up meta-analyses, consistently report increased circulating inflammatory markers and cytokines - including C-reactive protein (CRP), interleukin-6 (IL-6), tumor necrosis factor-α (TNF-α), interferon-γ (IFN-γ), and interleukin-1β (IL-1β) - and reduced levels of the anti-inflammatory cytokine interleukin-10 (IL-10) in patients with PTSD (Eraly et al., 2014; Passos et al., 2015; Yang & Jiang, 2020; Yuan et al., 2019). Furthermore, hypothesis-free approaches, including genome (Nievergelt et al., 2024; Stein et al., 2016) and epigenome-wide (Smith et al., 2011, 2020; Snijders et al., 2020; Uddin et al., 2010) association studies and transcriptomic profiling (Breen et al., 2018; Kuan et al., 2017, 2019; Pace et al., 2012), report immune and inflammatory pathways among the most dysregulated ones in PTSD.

Function and phenotype of immune cell subpopulations are determined by cell-specific metabolism (Buck et al., 2017; Hu et al., 2024). In the process known as metabolic reprogramming, immune cells adopt specific metabolic pathways/programs that enable them to carry out their effector functions (Buck et al., 2015, 2017; Kelly & O’Neill, 2015; Pearce et al., 2013). The most important metabolic pathways in immune cells include glycolysis, oxidative phosphorylation, fatty acid oxidation, fatty acid synthesis, the tricarboxylic acid (TCA) cycle, the pentose phosphate pathway, and amino acid metabolism (Hu et al., 2024; O’Neill et al., 2016). Despite extensive evidence of immune dysregulation in PTSD, no study has examined metabolic programming in immune cells from patients with PTSD or investigated its relationship to symptom severity.

In this exploratory case-control study, we investigated immunometabolic alterations in patients with PTSD across systemic, cellular, and transcriptional levels. Systemic inflammation and metabolic dysfunction were assessed through serum levels of IL-6, TNF-α, lactate, and growth differentiation factor-15 (GDF-15). We performed metabolic flow cytometry on peripheral blood mononuclear cells to quantify protein expression of key rate-limiting enzymes in major metabolic pathways (glycolysis, TCA cycle, pentose phosphate pathway, fatty acid synthesis, fatty acid oxidation, and adenosine triphosphate (ATP) synthesis). To capture transcriptional signatures of immune activation, we quantified gene expression of *TNF*, *NFKB1*, and *NR3C1*, representing key regulators of inflammatory and stress-response pathways relevant to PTSD. Finally, we explored associations between these immunometabolic markers and PTSD symptom severity.

## 2. Methods

### 2.1. Participants

This exploratory case-control study included peripheral blood mononuclear cells (PBMCs) and serum samples from two independent studies: The PTSD group (n = 34) consisted of a subgroup of participants diagnosed with PTSD according to DSM-5 criteria, with blood samples obtained prior to investigational medicinal product (IMP) administration in the THC-PTSD-trial (Ethik-Kommission des Landes Berlin, reference: 19/0384 – IV E 11) (Roepke et al., 2023). PTSD symptom severity was assessed using the Clinician-Administered PTSD Scale for DSM-5 (CAPS-5) (Weathers et al., 2018), the PTSD Checklist for DSM-5 (PCL-5) (Blevins et al., 2015) and comorbid depressive symptoms using the Montgomery–Åsberg Depression Rating Scale (MADRS) (Montgomery & Asberg, 1979). German validated versions of all instruments were administered. A non-clinical healthy control (HC) group (n = 32) was obtained from the study “Applying RdoĆs social processing domain to autism and social anxiety disorder: the role of neurobiology, social stress/support, and immunological markers” (RdoC-ASS/SAS; Ethikkommission Charité–Universitätsmedizin Berlin, reference: EA1/106/21). HCs were screened using a structured online self-report assessment based on DSM-5 criteria to exclude current and lifetime psychiatric disorders. Previous trauma exposure was assessed using items adapted from a DSM-5–based trauma history checklist (Life Events Checklist–5, LEC-5) (Weathers et al., 2013), depressive symptoms using Beck Depression Inventory-II (BDI-II) (Beck et al., 1996) and current PTSD symptom severity was assessed using PCL-5 (Blevins et al., 2015). Blood sampling, PBMCs isolation, serum processing, and long-term biobanking were performed according to identical standard operating procedures (SOPs, described in 2.2.1) according to identical protocol at Charité – Universitätsmedizin Berlin. All participants provided written informed consent in accordance with the principles outlined in the Declaration of Helsinki.

### 2.2. Immunometabolic analyses

To assess immunometabolic alterations associated with PTSD, we used a multimodal approach integrating metabolic flow cytometry to investigate key metabolic pathways across major immune cell subsets, serum analyses of immune and metabolic markers, and qPCR-based profiling of inflammatory and stress-related genes in purified immune cell subsets.

#### 2.2.1. Isolation and processing of peripheral blood mononuclear cells (PBMCs)

Peripheral blood mononuclear cells (PBMCs) were isolated from EDTA-anticoagulated blood using density gradient centrifugation (SepMate tubes with Ficoll-Paque PLUS or Biocoll). Following plasma removal, the PBMCs layer was collected, washed with phosphate-buffered saline (PBS) buffer, and treated with erythrocyte lysis buffer when necessary. After final washing, cells were resuspended in RPMI 1640 cell culture medium supplemented with 25% fetal bovine serum (FBS) Superior (Bio-Sell), counted using Trypan Blue exclusion, and cryopreserved in cell culture medium containing 10% dimethyl sulfoxide (DMSO) (Sigma-Aldrich). Samples were cooled to −80°C in a controlled-rate freezing container (Mr Frosty, Thermo Fisher Scientific) and after 24 hours transferred to liquid nitrogen for long-term storage at −196°C. For analyses, PBMCs were thawed and washed in prewarmed cell culture medium containing 10% FBS before downstream analyses.

#### 2.2.2. Serum analyses

Serum concentrations of IL-6, TNF-α, L-lactate, and GDF-15 were quantified using commercially available enzyme-linked immunosorbent assays (ELISA) (IL-6: Quantikine D6050B; TNF-α: Quantikine HSTA00E; GDF-15: Quantikine DGD150, R&D Systems; L-lactate: ab65330, Abcam) according to the manufacturers’ instructions. All samples were measured in duplicates, and optical density was recorded using a Tecan Infinite 200Pro plate reader. Concentrations were calculated from standard curves. All samples were analyzed on the same day to minimize inter-assay variability.

#### 2.2.3. Metabolic flow cytometry

To investigate immune cell metabolic programming in PTSD, we employed an adapted version of the Fluorescent Activated Cell Sorting (FACS)-based metabolic flow cytometry MetFlow assay (Ahl et al., 2020; Sattler et al., 2024). This method was optimized in our laboratory (Sattler et al., 2024) to enable simultaneous quantification of immune phenotype and metabolic enzyme expression at the single-cell level. We selected metabolic markers representing rate-limiting enzymes of major metabolic pathways in immune cells. Glucose transporter 1 (GLUT1) and hexokinase II (HKII) were included as indicators of glucose uptake and glycolytic activity, while lactate dehydrogenase (LDH) reflected the conversion of pyruvate to lactate during glycolysis. Glucose-6-phosphate dehydrogenase (G6PD) was measured as the key enzyme of the oxidative pentose phosphate pathway. To capture mitochondrial metabolism, isocitrate dehydrogenase 2 (IDH2) and ATP synthase subunit alpha 5 (ATP5A) were assessed as markers of the tricarboxylic acid (TCA) cycle and oxidative phosphorylation, respectively. Finally, carnitine palmitoyl transferase 1A (CPT1A) and acetyl-CoA carboxylase (ACAC) were analyzed as rate-limiting enzymes in fatty acid oxidation and fatty acid synthesis.

Cryopreserved PBMCs were thawed, washed, and stained using two antibody panels (Supplementary Table S1). The first panel targeted CD4+ and CD8+ T cells, including naïve and memory subsets defined by CD45RA and CCR7 expression, along with CD25 and CD127 used to distinguish regulatory T cells (Tregs). The second panel targeted non-T cell subsets, including B cells (CD19+), natural killer cells (CD56+), and monocytes (CD14+), together with HLA-DR, CD16 and CD11c for further phenotypic definition of cellular subsets. Cell viability was assessed using a Zombie Aqua viability dye (Biolegend). Samples were acquired on a BD LSRFortessa X-20 flow cytometer.

##### 2.2.3.1 Computational analysis of flow cytometry data

Compensation and initial data cleaning were performed in FlowJo (v10.8.1, BD Biosciences). Following gating of viable singlets, CD3+ T cells (Panel A) and total live cells (Panel B) were exported to the OMIQ analysis platform (https://www.omiq.ai/) for normalization (Van Gassen et al., 2020) and unsupervised clustering using the CITRUS (Cluster Identification, Characterization, and Regression) algorithm (Bruggner et al., 2014). For Panel A, CITRUS clustering was performed on 10.000 CD3+ T cells per sample based on surface marker expression to identify CD4+ and CD8+T cell subsets, followed by group level comparisons of the median fluorescence intensity (MFI) across the eight metabolic markers using SAM (Significance Analysis of Microarrays) model. For Panel B, CITRUS clustering was performed on 10.000 live cells per sample using surface markers to identify monocytes, B cells, and NK cells, and group comparison between HC and PTSD was conducted across the same eight metabolic markers. CITRUS analyses were repeated three times to verify the stability and reproducibility of clustering results. The most robust statistically significant clusters, corresponding to the major immune cell subsets, significant for FDR=0.01, were exported for downstream analyses. MFIs from these clusters were used for correlation with clinical variables (PCL-5 and MADRS scores). In addition, CITRUS results were exported for visualization and statistical validation of group-level differences in R (v4.2.3) (R Core Team, 2023), confirming the robustness of the findings.

#### 2.2.4. Gene expression in sorted immune cell subsets

To assess cell-specific transcriptional regulation of inflammatory and stress-related pathways, PBMCs were thawed, counted, and resuspended in buffer containing PBS, 2 mM ethylenediaminetetraacetic acid (EDTA), and 0.2% bovine serum albumin (BSA) (Miltenyi Biotec). Magnetic-activated cell sorting (MACS, Miltenyi Biotec) was used to isolate CD14+ monocytes, CD4+ T cells, and CD8+ T cells using antibody-conjugated magnetic microbeads. Sorted fractions were suspended in RNAprotect Cell Reagent (Qiagen) and stored at 4°C until RNA isolation. Sorting efficiency and purity were verified by flow cytometry (BD FACSCanto II) using fluorophore-conjugated antibodies against CD3, CD4, CD8, CD14, CD16, CD19, and CD56, and data were analysed with FlowJo (v10.10.0, BD Biosciences). Total RNA was isolated using the RNeasy Plus Mini Kit (Qiagen) with on-column DNase removal and QIAshredder homogenization. RNA quality and concentration were assessed by NanoDrop spectrophotometry. Complementary DNA (cDNA) was synthesised from 100–150 ng total RNA using the SuperScript IV First-Strand Synthesis System (Invitrogen). Quantitative PCR (qPCR) was performed on a StepOne™ Real-Time PCR System (Applied Biosystems) using TaqMan Gene Expression Assays (Applied Biosystems) for *NR3C1* (nuclear receptor subfamily 3 group C member 1, encoding glucocorticoid receptor, Hs00353740_m1), *NFKB1* (nuclear factor kappa B subunit 1, encoding NF-κB p50, Hs00765730_m1), and *TNF* (tumor necrosis factor, encoding TNF-α protein, Hs00174128_m1), with *TBP* (TATA-box binding protein, Hs00427620_m1) and *IPO8* (importin 8, Hs00183533_m1) as reference genes. All reactions were run in triplicate, and relative expression levels were calculated using the ΔCt method.

### 2.3. Statistical analysis

All statistical analyses and data visualizations were performed in R (v4.2.3). Group differences in serum marker concentrations and gene expression levels were assessed using Wilcoxon rank-sum tests on natural log-transformed serum concentrations or ΔCT values, respectively. Associations between metabolic flow cytometry markers and clinical variables, as well as between serum markers and clinical variables, were examined using Spearman’s rank correlation coefficients and corresponding two-tailed p-values using asymptotic t-distribution. Statistical significance was defined as p < 0.05.

## 3. Results

### 3.1. Sample characteristics

An overview of the sample characteristics is presented in Table 1. PTSD and HC groups did not differ significantly in age (mean ± SD: PTSD = 38.8 ± 12.3 years; HC = 34.1 ± 8.00 years, Welch t-test (57.09) = –1.85, p = 0.069) or gender distribution (61.8% vs. 62.5% female, χ² = 0, p = 0.95). Gender identity was self-reported as “male” or “female.” Sex assigned at birth was additionally recorded in the PTSD group (22 female, 12 male); one participant reported a gender identity different from their assigned sex. Body mass index (BMI) was significantly higher in the PTSD group compared to healthy controls (mean ± SD: 27.9 ± 7.6 vs. 24.2 ± 4.2 kg/m²; p = 0.018), consistent with prior literature reporting elevated rates of overweight and metabolic syndrome in PTSD patients (Bartoli et al., 2025; Suliman et al., 2016; Rosenbaum et al., 2015; Farr et al., 2015). Given the well-established links between PTSD and obesity as well as metabolic syndrome, BMI may represent both a comorbid metabolic condition and a potential mediator of systemic inflammation. Therefore, BMI was included as a covariate to determine whether immunometabolic alterations associated with PTSD persisted independently of overall adiposity. Age was included as an additional covariate due to its known influence on immune and metabolic function. The majority of PTSD patients were not taking any medication (24/34, 70.6%). Among those who were medicated, 5/34 (14.7%) were receiving non-psychiatric medication only, 3/34 (8.8%) psychiatric medication only, and 2/34 (5.9%) both non-psychiatric and psychiatric medications. One HC participant was taking psychiatric medication. A substantial proportion of HCs (24/32, 75.0%) reported exposure to at least one potentially traumatic event, which is consistent with reported prevalence rates of trauma exposure in the general population (Kessler et al., 2017). Due to occasional missing biomaterial or incomplete measurements (did not meet quality control thresholds or fell below assay detection limits), sample size varied across assays. A detailed overview of sample sizes per marker is provided in Supplementary Table S2.

**Table 1.** Sociodemographic and clinical characteristics of the study sample.

|  | HC | PTSD | p |
| --- | --- | --- | --- |
| Female, n (%) | 20 (62.5) | 21 (61.8) | 0.95 |
| Age, mean ± SD | 34.1 ± 8.00 | 38.8 ± 12.3 | 0.069 |
| BMI, mean ± SD | 24.2 ± 4.2 | 27.9 ± 7.6 | 0.018 |
| Any lifetime trauma exposure (LEC-5), n (%) | 24 (75.0) | 34 (100) |  |
| PCL-5 mean ± SD | 10.0 ± 11.2 | 51.1 ± 14.1 | < 0.001 |
| CAPS-5 mean ± SD | | 42.1 ± 11.9 | |
| MADRS mean ± SD | | 17.3 ± 6.8 | |
| BDI-II mean ± SD | 5.0 ± 6.3 | | |
| Medication: None / non-psychiatric / psychiatric / both | 31 / 0 / 1 / 0 | 24 / 5 / 3 / 2 |  |

Continuous variables were compared using Welch's t-tests; categorical variables were compared using $\chi^2$ tests. Symptom scales are reported descriptively. PCL-5: Posttraumatic Stress Disorder Checklist for DSM-5; CAPS-5: Clinician-Administered PTSD Scale for DSM-5; MADRS: Montgomery–Åsberg Depression Rating Scale; BDI-II: Beck Depression Inventory–Second Edition; BMI: Body Mass Index; LEC-5: Life Events Checklist for DSM-5.

### 3.2. Systemic inflammatory and metabolic markers in PTSD

To examine immunometabolic alterations on a systemic level, serum concentrations of pro-inflammatory and metabolic markers were compared between groups of PTSD and HC. Among the analyzed markers, IL-6 levels were significantly higher in the PTSD group (p = 0.005), whereas TNF-α, lactate, and GDF-15 did not differ significantly between groups. A slightly elevated, however not statistically different, lactate level was observed in PTSD (p = 0.08) (Figure 1). In models adjusting for age and BMI, the IL-6 group difference was attenuated and remained at a trend level (p = 0.057), indicating partial overlap with age- and BMI-related variance (Supplementary Figure 1A).

**Figure 1.**
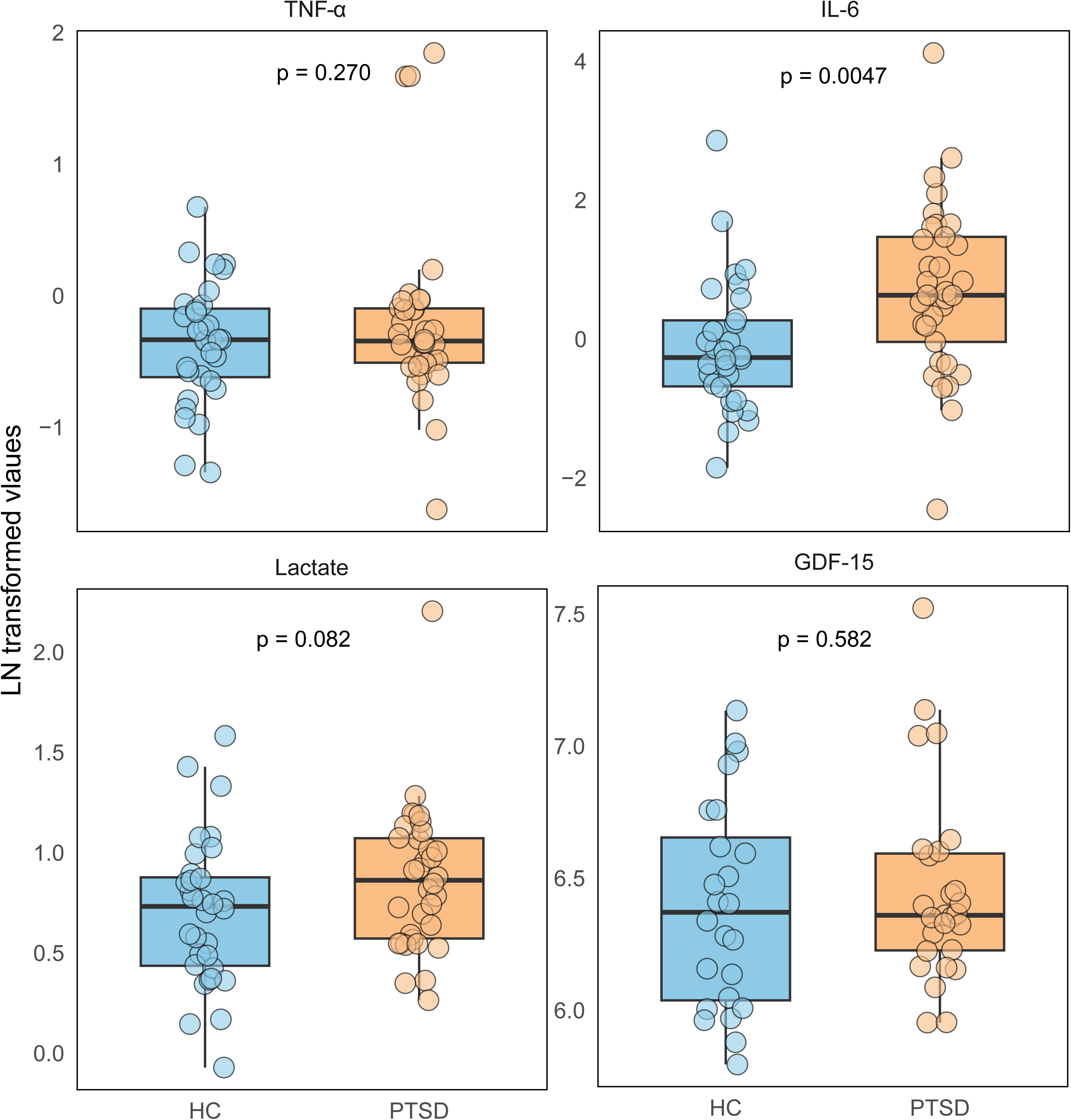
Systemic inflammatory and metabolic markers in PTSD. Boxplots show serum levels of TNF-α, IL-6, lactate, and GDF-15 in healthy controls (HC) and patients with post-traumatic stress disorder (PTSD). Data are shown as natural log (LN)-transformed values. TNF-α: Tumor necrosis factor-α; IL-6: Interleukin-6; GDF-15: Growth differentiation factor 15.

To explore the potential clinical relevance of these circulating markers, Spearman correlations were calculated between serum marker levels and CAPS-5 measured PTSD symptom severity within the PTSD group, revealing positive associations of GDF-15 and lactate with specific PTSD symptom domains as well as with depressive symptom severity (Supplementary Figure 2A).

Overall, these findings suggest an association between PTSD and systemic inflammation, reflected by higher IL-6 levels, in line with previous reports in the literature, although this association was attenuated after adjustment for age and BMI.

### 3.3. Immunometabolic alterations across immune cell subsets in PTSD

To explore cellular immunometabolism in PTSD, CITRUS analysis was applied to flow cytometry data to identify immune cell subsets exhibiting differential metabolic marker expression between groups of PTSD and HCs (Figure 2A–B).

**Figure 2.**
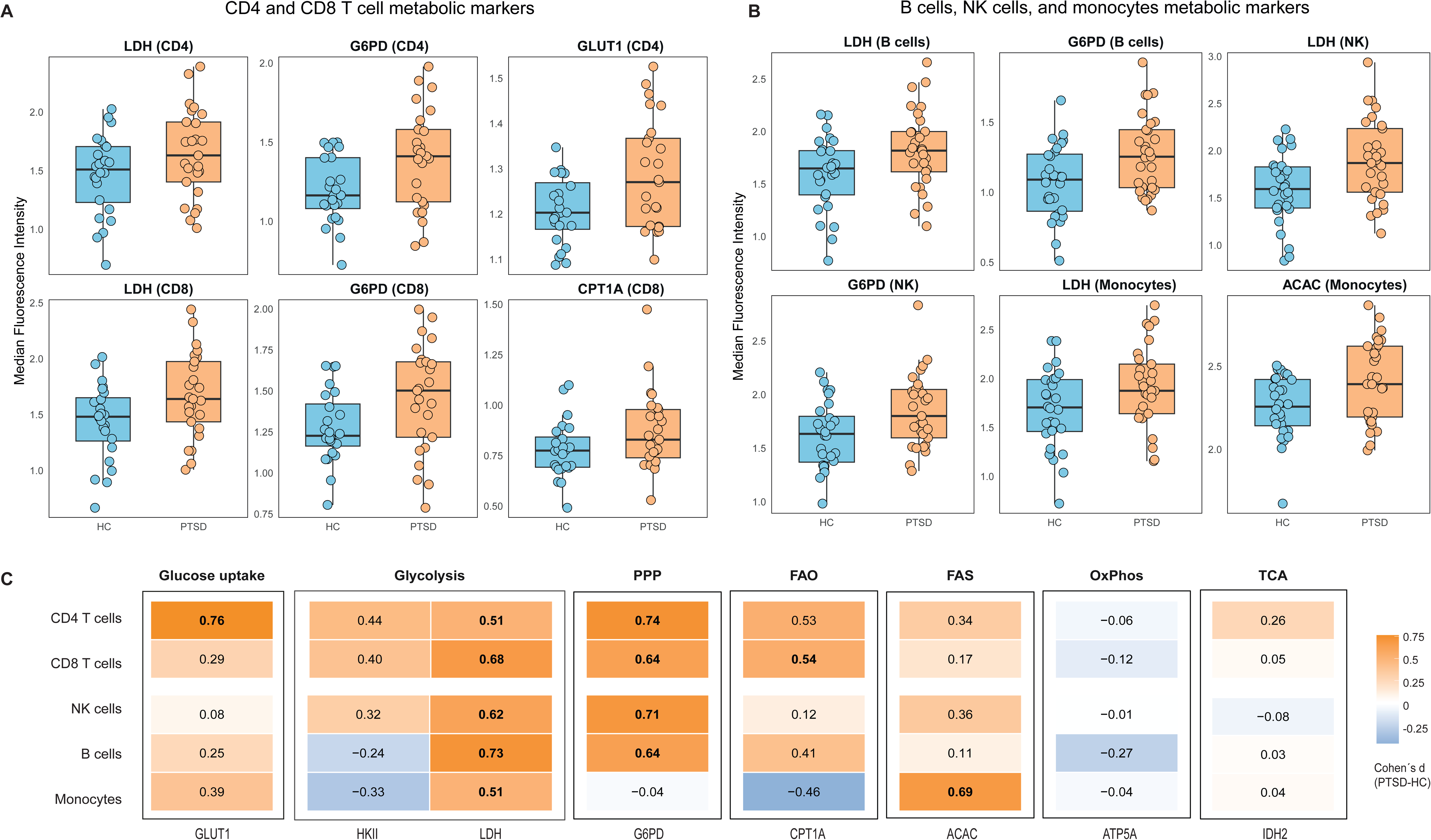
Immunometabolic reprogramming across adaptive and innate immune cell subsets in PTSD. Boxplots show metabolic markers identified by CITRUS (Cluster Identification, Characterization, and Regression) analysis as significantly differentially expressed between healthy controls (HC) and patients with post-traumatic stress disorder (PTSD). Boxplots (A–B) display median fluorescence intensity (MFI) of these markers across immune cell subsets: CD4+ and CD8+ T cells (A), B cells, natural killer (NK) cells, and monocytes (B). Heatmap (C) displays effect sizes (Coheńs d) for group differences in all metabolic markers across immune cell subsets. Effect sizes of markers identified by CITRUS as significantly different between groups, shown in boxplots A–B, are bold. GLUT1: Glucose transporter 1; HKII: Hexokinase II; LDH: Lactate dehydrogenase; G6PD: Glucose-6-phosphate dehydrogenase; CPT1A: Carnitine palmitoyl transferase 1A; ACAC: Acetyl-CoA carboxylase; IDH2: Isocitrate dehydrogenase 2; ATP5A: ATP synthase subunit alpha 5; PPP: Pentose Phosphate pathway; FAO: Fatty acid oxidation; FAS: Fatty acid synthesis; OxPhos: Oxidative phosphorylation; TCA: Tricarboxylic acid cycle.

Within the adaptive immune arm, CD4+ and CD8+ T cells (Figure 2A) from the PTSD group showed significantly increased expression of LDH and G6PD, indicating enhanced glycolytic and oxidative pentose phosphate pathway activity. In addition, CD4+ T cells exhibited significantly elevated levels of glucose transporter GLUT1, suggesting increased cellular glucose uptake capacity, while CD8+ T cells showed higher levels of CPT1a, reflecting enhanced engagement of fatty acid oxidation. CD19+ B cells (Figure 2B) also displayed significantly higher LDH and G6PD levels, reflecting a metabolic shift toward glycolysis and the oxidative pentose phosphate pathway. Among innate immune cells, CD14+ monocytes showed significantly increased LDH and ACAC expression, indicative of enhanced reliance on glycolysis and lipid biosynthesis, while CD56+ NK cells exhibited significantly elevated LDH and G6PD, reflecting a similar activation of glycolytic and pentose phosphate pathways (Figure 2B). Consistent with the clustering results, unadjusted standardized effect sizes were largely in the moderate range across glycolytic and pentose phosphate pathway markers, whereas markers related to oxidative phosphorylation and the TCA cycle showed minimal between-group differences (Figure 2C).

To determine whether these immunometabolic alterations were independent of adiposity and age, we performed linear regression analyses adjusting for BMI and age. Several metabolic markers remained elevated in PTSD following covariate adjustments, including GLUT1 in CD4+ T, LDH and G6PD in B cells, LDH and G6PD in NK cells, and ACAC in monocytes (Supplementary Figure 3A). For other markers, adjustment reduced the magnitude of the group difference, although effects remained directionally consistent with the unadjusted analyses. Adjusted effect sizes were in the small-to-moderate range, suggesting that the observed alterations are not fully explained by differences in BMI or age, and may reflect PTSD-related immunometabolic reprogramming (Supplementary Figure 3B).

To explore the potential clinical relevance of these immunometabolic alterations, Spearman correlations between marker expression and CAPS-5 measured PTSD symptom severity were calculated within the PTSD group, revealing significant associations particularly for LDH and G6PD (Supplementary Figure 2B).

Together, these findings indicate widespread immunometabolic alterations in PTSD, predominantly within the glycolytic and pentose phosphate pathways, typical of immune cell activation across adaptive and innate immune subsets.

### 3.3. Transcriptional markers of inflammatory and stress-related pathways in PTSD

To assess inflammatory and stress-related signaling at the transcriptional level, gene expression of *NR3C1*, *NFKB1*, and *TNF* was quantified in MACS-sorted immune cell subsets in groups of PTSD and HC (Figure 3A–C). In CD4+ T cells (Figure 3A), *TNF* expressions were elevated in the PTSD group (*p* = 0.046). After adjustment for age and BMI, the group difference was attenuated and no longer statistically significant (*p* = 0.149) (Supplementary Figure 1B). No significant group differences were observed in CD8+ T cells (Figure 3B) or CD14+ monocytes (Figure 3C) for any of the analyzed genes.

**Figure 3.**
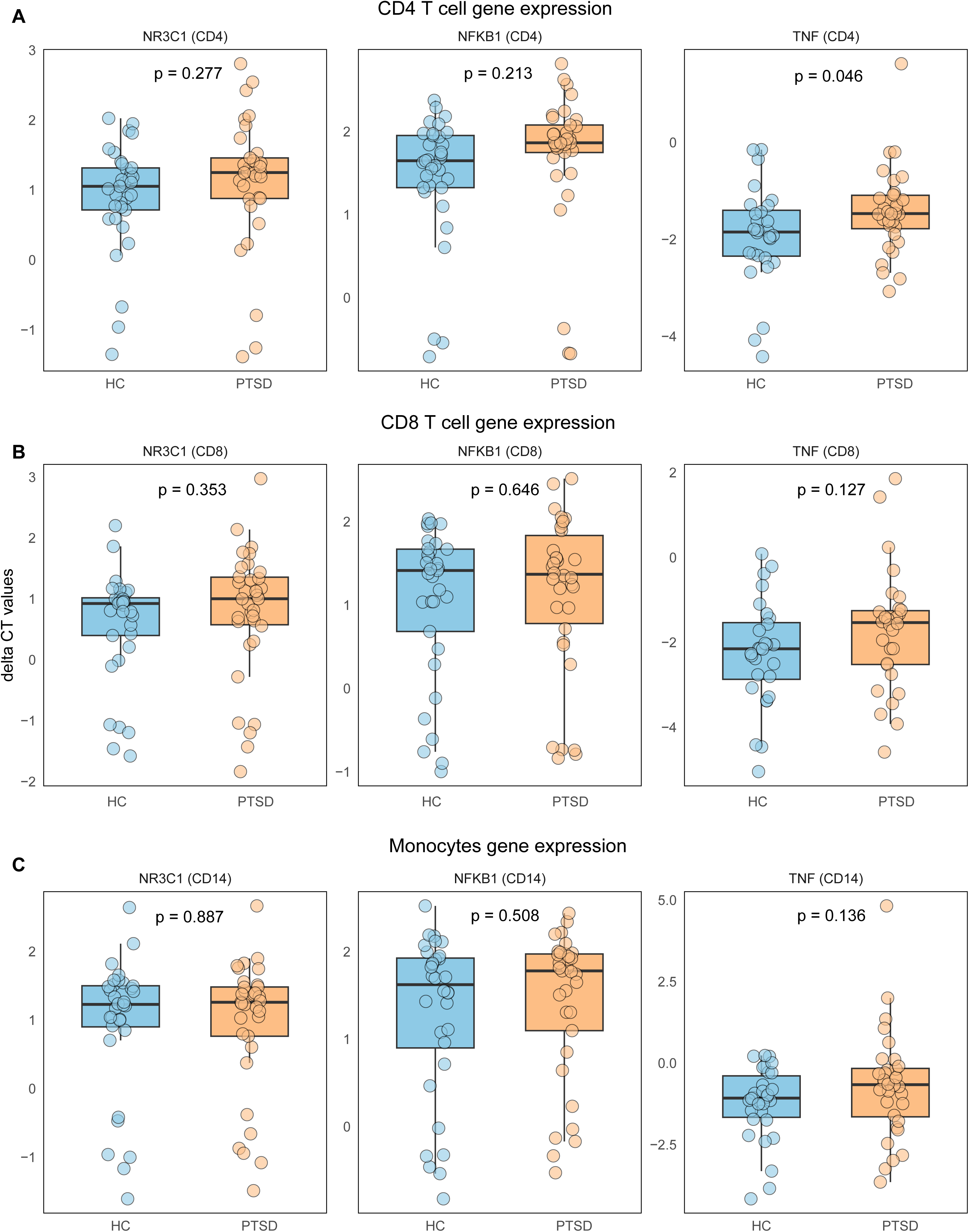
Transcriptional markers of inflammatory and glucocorticoid signaling in immune cell subsets in PTSD. Boxplots show relative mRNA levels (ΔCt values) of *NR3C1*, *NFKB1,* and *TNF* in sorted immune cell subsets from healthy controls (HC) and patients with post-traumatic stress disorder (PTSD). (A) CD4+ T cells, (B) CD8+ T cells, and (C) CD14+ monocytes. *NR3C1*: Nuclear receptor subfamily 3 group C member 1, encoding glucocorticoid receptor; *NFKB1*: Nuclear factor kappa B subunit 1, encoding NF-κB p50; *TNF*: Tumor necrosis factor, encoding TNF-α protein.

Taken together, these results point to largely comparable expression profiles of inflammation- and stress-related genes across major immune cell subsets in PTSD and HCs.

## 4. Discussion

In this exploratory case-control study, we investigated immunometabolic alterations in patients with PTSD on systemic, cellular, and transcriptional levels. To our knowledge, this is the first study to investigate immune cell metabolism in PTSD. Immune cell metabolic profile revealed a consistent increase in LDH and G6PD expression in circulating immune cells (Figure 1A–B, C), suggesting enhanced glycolytic and oxidative pentose phosphate pathway activity. These pathways are canonical features of the metabolic reprogramming that accompanies immune activation, where cells transition from a quiescent, oxidative phosphorylation-dependent state toward a pro-inflammatory effector phenotype supported by rapid energy generation through glycolysis (Buck et al., 2017; Chapman et al., 2020; Hu et al., 2024; O’Neill et al., 2016). In our study, this metabolic shift was evident across major cell subsets within both adaptive and innate arms of the immune system. While preliminary, these findings suggest that altered cellular metabolism may be a feature of PTSD-related immune dysregulation.

Systemic findings were broadly consistent with this activated immune phenotype. Patients with PTSD exhibited elevated circulating IL-6 (Figure 1A), one of the most consistently replicated inflammatory cytokines in PTSD (Passos et al., 2015). Although the group difference was attenuated after adjustment for age and BMI, the direction and magnitude of the effect remained comparable, suggesting that demographic and metabolic factors account for part, but not all, of the observed elevation. This is consistent with the notion that systemic inflammation in PTSD may be intertwined with broader metabolic alterations. Serum lactate levels showed a slight, trend-level increase (p = 0.08) (Figure 1A), which while not statistically significant may be indicative of enhanced glycolytic activity, as lactate is generated through LDH-mediated conversion of pyruvate and reflects increased glycolytic flux. (Hu et al., 2024; O’Neill et al., 2016). This pattern aligns with metabolomic findings in PTSD, where elevated lactate points to enhanced dependence on glycolytic pathways and mitochondrial dysfunction (Mellon et al., 2018, 2019). Given that increased BMI may represent both a comorbid condition and a potential downstream consequence of PTSD-related behavioral and metabolic alterations, adjusted models should be interpreted as estimating PTSD-related effects independent of adiposity, rather than implying that BMI is unrelated to PTSD pathophysiology. Taken together, the pattern across systemic and cellular levels suggests that while circulating inflammatory markers may partly overlap with broader metabolic and demographic factors, immune cell metabolic alterations appeared comparatively more consistent across analytical models. This observation raises the possibility that cellular metabolic alterations may represent a more stable feature of PTSD-related immune dysregulation, rather than merely reflecting downstream effects of systemic inflammation or adiposity.

Beyond group-level differences, metabolic marker expression was associated with overall PTSD symptom severity (total CAPS-5 score) as well as with individual symptom domains (Avoidance and Arousal & Reactivity). Further mechanistic and longitudinal studies are needed to disentangle this relationship and to determine whether distinct immunometabolic phenotypes underly different PTSD symptom profiles.

Several limitations should be acknowledged. The sample size was relatively small and may have limited statistical power to detect additional group differences or subtle effects. Given the cross-sectional design, causal inferences cannot be drawn, and it remains unknown whether these metabolic alterations represent pre-existing vulnerability, consequences of chronic stress, or markers of disease severity. Longitudinal studies are needed to determine whether symptom improvement or treatment response is accompanied by normalization of metabolic markers. Additionally, we could not control for menstrual cycle phase, physical activity, sleep, or diet. Furthermore, metabolic assessments were limited to a set of eight metabolic markers and future work should incorporate higher-resolution metabolic assays, mitochondrial functional measures, and deeper investigation of lactate metabolism to more precisely dissect the mechanistic basis of immunometabolic alterations in PTSD.

In summary, these findings suggest that PTSD may be characterized by immunometabolic alterations with potential translational relevance, as targeting dysregulated immune and metabolic pathways could represent a promising avenue for future symptom-focused interventions.

## Supporting information

Supplementary material Brasanac PTSD Immunometabolism

## Data availability

Data will be made available on request.

## Author contributions

**Jelena Brasanac**: Conceptualization, Methodology, Investigation, Formal Analysis, Data Curation, Visualization, Writing – Original Draft, Writing – Review & Editing. **Linda El-Ahmad:** Investigation, Resources, Data Curation, Writing – Review & Editing. **Emma Mollerup:** Investigation, Data Curation, Writing – Review & Editing. **Stefanie Gamradt:** Investigation, Methodology, Writing – Review & Editing. **Luisa Gruenberg:** Investigation, Resources, Writing – Review & Editing. **Daria Shyshko:** Investigation, Writing – Review & Editing. **Victoria Stiglbauer:** Investigation, Methodology, Writing – Review & Editing. **Kim Zimbalski:** Investigation, Writing – Review & Editing. **Nikola Schoofs:** Resources, Writing – Review & Editing. **Kathlen Priebe:** Resources, Writing – Review & Editing. **Felix Wülfing:** Resources, Writing – Review & Editing. **Simon Guendelman:** Resources, Data Curation, Funding Acquisition, Writing – Review & Editing. **Tolou Maslahati:** Resources, Writing – Review & Editing. **Stefanie Koglin:** Resources, Writing – Review & Editing. **Christian Otte:** Resources, Writing – Review & Editing. **Isabel Dziobek:** Resources, Funding Acquisition, Writing – Review & Editing. **Stefan Roepke**: Resources, Funding Acquisition, Writing – Review & Editing. **Stefan M. Gold:** Conceptualization, Methodology, Supervision, Resources, Funding Acquisition, Writing – Review & Editing.

