## Supplementary material Brasanac PTSD Immunometabolism for "Immunometabolic Alterations in Post-Traumatic Stress Disorder"

**Supplementary Table S1.** Antibody panels for metabolic flow cytometry

| **Panel A – T cells** | **Panel B – non T cells** |
|---|---|
| Surface markers | Surface markers |
| CD3 | CD3 |
| CD4 | CD11c |
| CD8 | HLA-DR |
| CD45RA | CD16 |
| CCR7 | CD14 |
| CD25 | CD56 |
| CD127 | CD19 |
| Zombie Aqua | Zombie Aqua |
| Metabolic markers | Metabolic markers |
| GLUT1 | GLUT1 |
| HKII | HKII |
| G6PD | G6PD |
| LDH | LDH |
| CPT1A | CPT1A |
| ACAC | ACAC |
| IDH2 | IDH2 |
| ATP5A | ATP5A |

**Supplementary Table S2.** Summary of assay-specific sample numbers

| Marker | N (PTSD) | N (HC) |
|---|---|---|
| CD4+ T cells NR3C1 | 33 | 32 |
| CD4+ T cells NFKB1 | 33 | 32 |
| CD4+ T cells TNF | 32 | 27 |
| CD8+ T cells NR3C1 | 34 | 32 |
| CD8+ T cells NFKB1 | 34 | 32 |
| CD8+ T cells TNF | 30 | 27 |
| Monocytes NR3C1 | 33 | 31 |
| Monocytes NFKB1 | 33 | 31 |
| Monocytes TNF | 32 | 29 |
| TNF-$\alpha$ | 34 | 32 |
| IL-6 | 33 | 30 |
| GDF-15 | 27 | 24 |
| Lactate | 34 | 32 |
| LDH CD4+ T cells | 25 | 24 |
| G6PD CD4+ T cells | 25 | 24 |
| GLUT1 CD4+ T cells | 25 | 24 |
| LDH CD8+ T cells | 25 | 24 |
| G6PD CD8+ T cells | 25 | 24 |
| CPT1A CD8+ T cells | 25 | 24 |
| LDH B cells | 29 | 28 |
| G6PD B cells | 29 | 28 |
| LDH NK cells | 29 | 28 |
| G6PD NK cells | 29 | 28 |
| LDH Monocytes | 29 | 28 |
| ACAC Monocytes | 29 | 28 |

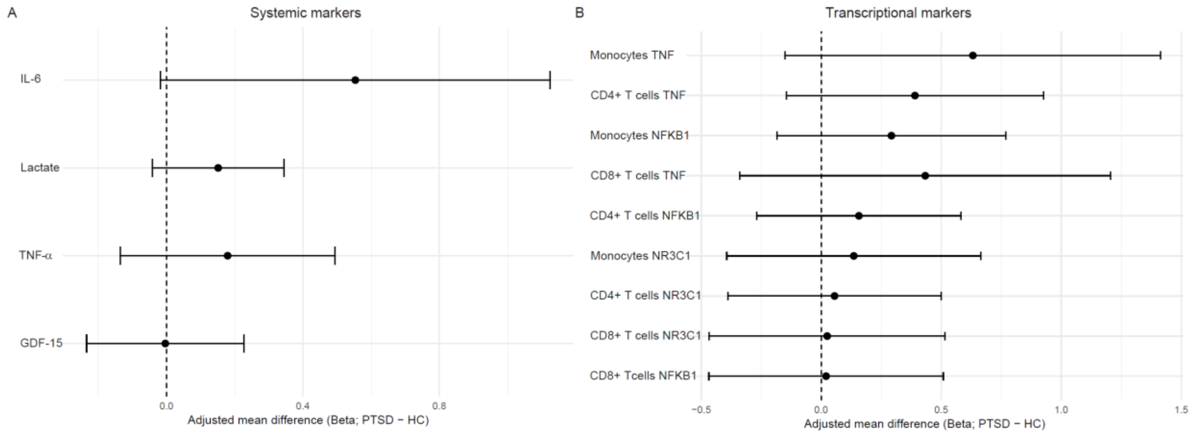


**Supplementary Figure 1. Age- and BMI-adjusted group differences in systemic and transcriptional markers.** Forest plots showing (A) adjusted mean differences (β) for circulating systemic markers concentrations and (B) adjusted mean differences (β) for immune cell-specific gene expression levels in post-traumatic stress disorder (PTSD) compared to healthy controls (HC). Estimates were derived from linear regression models including group (PTSD vs HC), age, and body mass index (BMI). Points represent estimated group differences (PTSD − HC), and horizontal lines indicate 95% confidence intervals. The dashed vertical line denotes no group difference (β = 0); positive values indicate higher marker levels or gene expression in PTSD. IL-6: Interleukin-6; TNF-α: Tumor necrosis factor alpha; GDF-15: Growth differentiation factor 15; NR3C1: Glucocorticoid receptor; TNF: Tumor necrosis factor; NFKB1: Nuclear factor kappa B subunit 1.

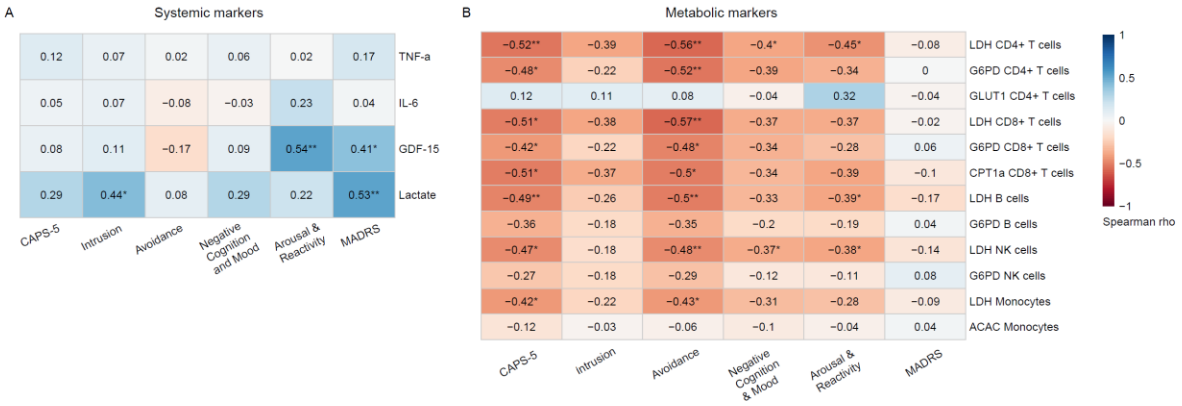


**Supplementary Figure 2. Associations between systemic and metabolic markers with symptom severity.** Heatmap (A) showing Spearman correlation coefficients (ρ) between systemic marker levels and symptom severity in post-traumatic stress disorder (PTSD) group, including total and subscale scores of the

Clinician Administered PTSD Scale 5 (CAPS-5) and total scores of the Montgomery–Åsberg Depression Rating Scale (MADRS). Heatmap (B) shows Spearman correlation coefficients (ρ) between metabolic marker expression (median fluorescence intensity) and symptom severity within the PTSD group, including total and subscale scores of the CAPS-5 and total scores of MADRS. Significant correlations (* $p < 0.05$, ** $p < 0.01$) are indicated by an asterisk. TNF-α: Tumor necrosis factor-α; IL-6: Interleukin-6; GDF-15: Growth differentiation factor 15. GLUT1: Glucose transporter 1; HKII: Hexokinase II; LDH: Lactate dehydrogenase; G6PD: Glucose-6-phosphate dehydrogenase; CPT1A: Carnitine palmitoyl transferase 1A; ACAC: Acetyl-CoA carboxylase; IDH2: Isocitrate dehydrogenase 2; ATP5A: ATP synthase subunit alpha 5; PPP: Pentose Phosphate pathway; FAO: Fatty acid oxidation; FAS: Fatty acid synthesis; OxPhos: Oxidative phosphorylation; TCA: Tricarboxylic acid cycle.

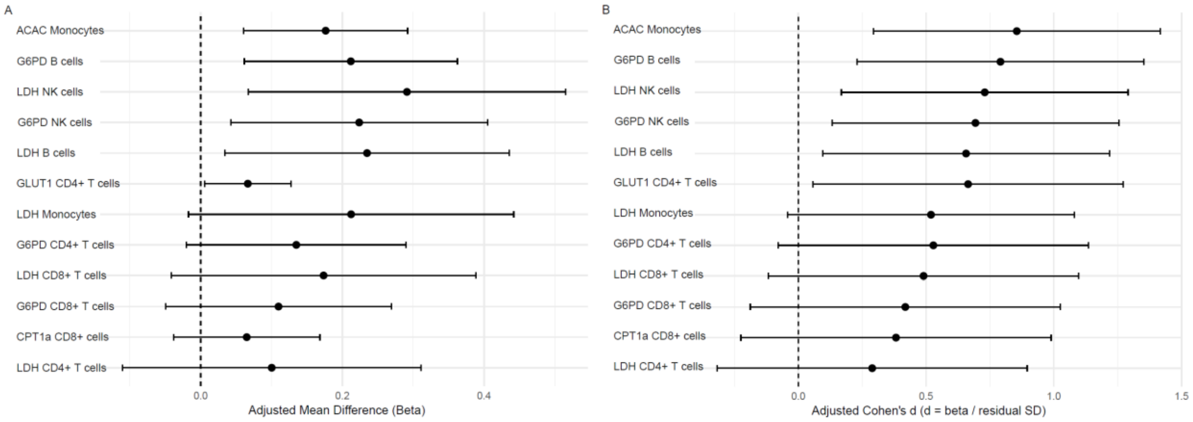


**Supplementary Figure 3. Age- and BMI-adjusted group differences in metabolic markers.** Forest plots showing (A) adjusted mean differences (β) and (B) adjusted Cohen's d values for metabolic marker expression across immune cell subsets in post-traumatic stress disorder (PTSD) compared to healthy controls (HC). Estimates were derived from linear regression models including group (PTSD vs HC), age, and body mass index (BMI). Adjusted Cohen's d was calculated as β divided by the residual standard deviation of the fitted model. Points represent estimated group differences (PTSD − HC), and horizontal lines indicate 95% confidence intervals. The dashed vertical line denotes no group difference (β or d = 0); positive values indicate higher marker expression in PTSD. GLUT1: Glucose transporter 1; LDH: Lactate dehydrogenase; G6PD: Glucose-6-phosphate dehydrogenase; CPT1A: Carnitine palmitoyl transferase 1A; ACAC: Acetyl-CoA carboxylase.
